# A Pilot Study of the Combination of 5-Azacitidine and All-trans Retinoic Acid in Biochemically Recurrent Prostate Cancer

**DOI:** 10.1101/2025.08.11.25333316

**Authors:** Vaibhav G. Patel, Deepak K. Singh, Himanshu Joshi, Nyima Sherpa, Bobby Liaw, Che-Kai Tsao, Matthew D. Galsky, Lindsay Diamond, Melisa Lopez-Anton, Maria Soledad Sosa, Julio A. Aguirre-Ghiso, William K. Oh

## Abstract

**Translational Relevance:** Despite definitive local therapy, some men with prostate cancer develop biochemical recurrence (BCR-PCa) and progress to metastatic disease. Current standard-of-care, androgen receptor pathway inhibitors (ARPIs) with or without androgen deprivation therapy (ADT) carry substantial long-term morbidity. This trial tested a novel, non-hormonal approach of 5-azacitidine (AZA) plus all-trans retinoic acid (ATRA) to induce tumor cell dormancy via epigenetic reprogramming. The regimen was well tolerated, with manageable toxicities, and showed preliminary signals of delayed PSA progression and prolonged PSA doubling time in some patients, suggesting dormancy induction. One patient achieved durable disease stabilization with ATRA maintenance. These first-in-human findings indicate that epigenetic reprogramming may modulate dormancy in BCR-PCa, offering a potential strategy to delay or minimize ADT use and its toxicities. This approach highlights the translational potential of preventing or delaying overt metastases by activating dormancy pathways in early recurrent prostate cancer.

**Purpose:** Biochemical recurrence (BCR) after definitive local therapy remains a major clinical challenge in prostate cancer (PCa), with heterogeneous disease trajectories and few established strategies to delay further progression without prolonged androgen deprivation. This pilot study evaluated the combination of 5-azacitidine (AZA) and all-trans retinoic acid (ATRA) to induce tumor dormancy and delay clinical progression in patients with BCR.

**Experimental Design:** In a prospective, open-label, randomized, single-institution pilot trial, patients with BCR of PCa and no recent hormonal or definitive therapy received low-dose AZA and sequential ATRA. The co-primary endpoints were changes in prostate-specific antigen doubling time (PSADT) and time to next treatment (TTNT). Safety and biomarker analyses, including bone morphogenetic protein (BMP) signaling and dormancy marker NR2F1 in circulating tumor cells (CTCs), were evaluated to investigate treatment effects on minimal residual disease dormancy.

**Results:** Fourteen patients were enrolled. Treatment resulted in an increase in median PSADT from 2.45 to 4.56 months. The median TTNT was 9.6 months, with 28.6% of patients experiencing TTNT over 12 months. No new safety signals were identified; adverse events were consistent with those expected for AZA and ATRA. Analysis of circulating BMP4 and BMP7 suggested that higher BMP4 levels may correlate with treatment response. Notably, all patients achieved testosterone recovery post-treatment, likely reflecting the avoidance of ongoing androgen deprivation. Across the cohort, treatment with AZA+ATRA led to a reduction in total CTC numbers and an apparent increase in the fraction of NR2F1-positive CTCs in responders, although the small cohort size limited statistical testing.

**Conclusions:** The combination of AZA and ATRA was feasible and prolonged PSA kinetics in a subset of patients with BCR of PCa, with a favorable safety profile. This epigenetic approach promoting tumor dormancy presents a potential strategy to defer progression and delay the need for continuous hormonal suppression. Larger studies are warranted to validate these findings and further explore biomarkers predictive of clinical benefit.

## Introduction

Prostate cancer (PCa) remains a leading cause of cancer-related morbidity and mortality worldwide. While definitive therapies such as radical prostatectomy or radiation can cure localized disease, up to half of the patients develop biochemical recurrence (BCR), defined as a rising prostate-specific antigen (PSA) level in the absence of radiographic metastases (1,2). This clinical state is heterogeneous, where some patients progress rapidly to overt metastases, while others experience prolonged periods of indolence. Androgen ablation (androgen deprivation therapy, ADT) in prostate cancer frequently induces a tumor dormancy-like phenotype, characterized by growth arrest and prolonged asymptomatic periods before potential relapse. This effect is mediated by the inhibition of androgen receptor signaling, which initially suppresses tumor proliferation but often leads to the survival of residual cancer cells in a quiescent or dormant state. This may also be due to existing cues (i.e., TGFβ2, BMP7) in the bone marrow that induce dormancy of prostate disseminated tumor cells (DTCs) that predate the treatment or persist in response to androgen-ablative therapies (3–7). Importantly, patients with no evidence of disease for over a decade more commonly carried DTCs in bone marrow at the time of surgery that expressed dormancy markers than those in an advanced state, where these markers were downregulated (8,9). A few of these dormancy markers indicated that retinoic acid signaling was being reinstated, including RARβ and NR2F1 in dormant PCa DTCs. Additionally, hormonal ablation also appears to induce dormancy markers such as NR2F1 (9,10). These data highlight the critical role of tumor dormancy in the progression of disease (11).

As mentioned above, tumor cell dormancy is regulated by intrinsic cellular programs and microenvironmental cues (4,8,9,11–13). Dormant DTCs can evade immune surveillance and are resistant to therapies targeting proliferating cells, serving as a reservoir for eventual metastatic outgrowth (14,15). Epigenetic mechanisms, particularly DNA methylation and chromatin remodeling, have been implicated in regulating dormancy (9,16). In preclinical models, inducing a dormancy-associated epigenetic state using a combination of the DNMT1 inhibitor 5-azacytidine / decitabine and all trans retinoic acid (ATRA, a differentiation agent) delayed metastatic progression and reprogrammed various cancer cell types, including prostate cancer cells, to express dormancy-regulating genes, offering a therapeutic strategy distinct from conventional cytotoxic or hormone-based approaches (9,17). The sequence of AZA followed by ATRA was implemented from the experimental studies showing that the AZA step renders cancer cells more responsive to ATRA in the second step compared to each drug alone (9).

AZA was tested in Phase II trials of patients with PCa. A phase II trial in 26 patients with castration-resistant prostate cancer (CRPC) and PSA doubling time (PSADT) <3 months used a dose of 75 mg/m^2^ subcutaneous on days 1-5, administered every 28 days (18). The primary endpoint was the prolongation of PSADT by more than 3 months, which was noted in 19 patients (55%). PSA declines were noted in 14 of 36 evaluable patients. Median PSADT was 2.8 months after treatment, compared with 1.5 months prior to starting (p<0.01), but at these doses, toxicity was notable. ATRA was also studied in Phase II trials; Trump et al. administered ATRA at 50 mg/m² TID for 14 days every 3 weeks to 17 CRPC patients and observed no clinical responses (19). However, a trial of ATRA 50 mg/m² daily, combined with cis-retinoic acid and IFN, showed that four patients with hormone-sensitive PCa experienced stabilization in PSA kinetics (20). However, no trial to date has combined both drugs, which we hypothesize will act synergistically for prolonged periods. Our experimental data supported that low-dose 5-Azacitidine (AZA) combined in sequence with all-trans retinoic acid (ATRA) may synergistically induce a dormant state by reactivating differentiation pathways and suppressing proliferative reprogramming. In prostate cancer, these agents may target DTCs and delay metastasis without requiring prolonged androgen deprivation.

To this end, we conducted a prospective, open-label, randomized pilot study (NCT03572387) to evaluate the safety and anti-metastatic activity of epigenetic reprogramming therapy with AZA + ATRA in men with BCR-PCa.

## Patients and Methods

### Study Design and Patient Population

This study was a prospective, open-label, randomized, single-institution pilot study designed to evaluate the effects of AZA and ATRA on disease progression in patients with biochemically recurrent prostate cancer. The study was conducted in accordance with the Declaration of Helsinki and Good Clinical Practice guidelines. Institutional Review Board (IRB) approval was obtained at the Icahn School of Medicine at Mount Sinai, and written informed consent was required from all patients prior to study enrollment.

Eligible patients are men with histologically confirmed prostate adenocarcinoma who have undergone prior radical prostatectomy and/or radiation therapy, with biochemical recurrence defined as a rising PSA level with PSADT ≤ 10 months, have an indication for androgen deprivation therapy (ADT), and have baseline testosterone levels > 50 ng/dl. Participants must have no radiographic evidence of metastatic disease on conventional imaging (bone scan and CT/MRI), except for regional metastasis where salvage radiation therapy is not an option. Participants who received stereotactic body radiation therapy (SBRT) for oligometastatic lesion(s) with no confirmed active radiographic disease, as determined by the treating physician, were allowed on the study.

Patients who received ADT and/or other anticancer therapies within 3 months prior to randomization, and those who underwent radiotherapy or surgery within 4 weeks prior to randomization, were excluded from the study.

### Randomization and Treatment

All enrolled patients were randomized in a 1:1 fashion to either the “early” or “delayed” treatment with 5-AZA+ATRA. Patients in both arms first received a single 7.5 mg dose of leuprolide acetate at baseline to normalize PSA kinetics prior to randomization. After the 4-week course of leuprolide, the “early treatment” group received 5-AZA + ATRA for 3 cycles, followed by no treatment for 3 cycles. The “delayed treatment” group received no treatment for 3 cycles, followed by 3 cycles of 5-AZA + ATRA. 5-AZA 40 mg/m^2^ was administered subcutaneously on Days 1-5 of each 28-day cycle, and ATRA 45 mg/m^2^ was administered orally, divided into two doses on Days 3-7 of each 28-day cycle. Patients continued to receive treatment for the full 3-cycle duration, unless there was evidence of radiographic progression, an unacceptable adverse event, or the patient or physician decided to discontinue the regimen. After the treatment phase (early = 5-AZA+ATRA → observation, delayed = observation → 5-AZA+ATRA), patients were continued to follow up for up to 24 months from day 1 of the study, unless there was evidence of radiographic progression and/or initiation of other cancer-directed therapies.

### Endpoints and Assessments

The primary endpoint of the study was the disease progression-free rate at 12 weeks after completing the treatment phase. Disease progression was defined as a composite of confirmed PSA or radiographic progression, or initiation of other cancer-directed therapies, or death related to disease or treatment. Key secondary endpoints were safety and tolerability, change in PSADT at 12 weeks after completion of the treatment phase, and time to next treatment (TTNT).

Details regarding baseline assessments, efficacy assessments, and adverse events are provided in **Table S1**. Baseline evaluations were performed within 4 weeks before the start of trial treatment, except for radiographic evaluation, which was allowed within 2 months of study initiation. PSA progression was defined by the Prostate Cancer Working Group 3 (PCWG3) criteria.(21) The baseline PSA was defined as the serum PSA level measured prior to the initiation of leuprolide on study. PSADT was calculated using the MSK nomogram (https://www.mskcc.org/nomograms/prostate/psa_doubling_time). TTNT was calculated from the receipt of leuprolide in the study to the initiation of other cancer-directed therapies. Radiographic progression was evaluated using the Response Evaluation Criteria in Solid Tumors (RECIST) version 1.1 and the PCWG3 criteria, with CT/MRI and whole-body radionuclide bone scans, respectively.(21,22) Adverse events were graded according to the Common Terminology Criteria for Adverse Events (CTCAE) version 4.03.

### Cytokine Analysis

Ten ml of whole blood drawn from patients was collected in EDTA-treated tubes and processed within 2 hours of collection. Whole blood samples were centrifuged at 1000g for 10 minutes at 4°C to separate plasma from blood cells. The upper plasma layer was transferred to a new centrifuge tube and further analyzed to detect the levels of BMP4 and BMP7. ELISA was performed using the kits (BMP4, cat # DY314, R&D) and (BMP7, cat # DY354, R&D), following the manufacturer’s protocol.

### Circulating Tumor Cells (CTCs)

CTCs isolated from the peripheral blood of the patients were fixed in 4% paraformaldehyde and attached to glass slides using the cytospin method. Cytospin slides were washed twice with 1X PBS for 10 minutes each, followed by a 1-hour blocking step with 3% BSA (Fisher Bioreagents, cat # BP1605) and 5% normal goat serum (Gibco, cat # PCN5000) in PBS at room temperature. Primary antibodies (pan-cytokeratin, abcam cat # ab27988; CD45, Ray Biotech cat # 119-17121; and NR2F1, abcam cat # ab181137) were incubated overnight at 4°C, followed by washing (PBS, 3 x 10 minutes) and incubation with Alexa-conjugated secondary antibodies (Invitrogen, cat # A-11001, cat # A-11077, cat # A-21245, 1:1000) at room temperature for 1-2 hours in the dark. Slides were washed (PBS, 3 x 10 minutes) and mounted with Prolong Gold antifade mounting medium with DAPI. Slides were scanned using a P250 Flash III scanner. Cells positive for DAPI, pan-CK, and negative for CD45 were considered CTCs.

### Statistical Analysis

Analysis was performed using custom scripts in the R statistical language (version 4.2.3). Continuous variables were compared using a t-test, and categorical variables were compared using a chi-square test for the descriptive analysis. The comparison of continuous variables was conducted using the Wilcoxon rank sum test, and signed rank tests were used in the case of nonparametric data for two-sample and one-sample cases, respectively. P < 0.05 was interpreted as statistically significant. The R package ‘swim plot’ was used to generate a Swimmer plot.

## Results

### Patients

Between August 2018 and November 2021, a total of 14 patients were enrolled and underwent randomization, with 6 assigned to the “early” arm and 8 assigned to the “delayed” arm. The data cut-off date was March 06, 2023. The baseline characteristics are listed in **Table 1**. The mean age was 66.4 (range, 52-76) years, mean PSA levels of 4.75 ng/ml, and mean PSADT of 2.89 months. In the study population, 57.1% had a Gleason score ≥ 8, and prior treatment consisted of RT + ADT, surgery + ADT, surgery + RT, surgery + RT + ADT in 3 (21.4%), 3 (21.4%), 3 (21.4%), and 5 (35.7%) patients, respectively. Additionally, 42.9% of patients received prior RT to oligometastatic lesion(s) with the absence of active radiographic disease prior to study enrollment.

**Table 1.** Baseline Patient Characteristics.

|  | **Delayed**<br>**(N=8)** | **Early**<br>**(N=6)** | **Overall**<br>**(N=14)** |
| --- | --- | --- | --- |
| **Age** |  |  |  |
| Mean (SD) | 62.3 (6.80) | 71.8 (5.04) | 66.4 (7.64) |
| **Race and ethnicity** |  |  |  |
| Hispanic White | 1 (12.5%) | 0 (0%) | 1 (7.1%) |
| Non-Hispanic White | 6 (75.0%) | 5 (83.3%) | 11 (78.6%) |
| Others | 1 (12.5%) | 1 (16.7%) | 2 (14.3%) |
| **Gleason Score** |  |  |  |
| <=7 | 5 (62.5%) | 0 (0%) | 5 (35.7%) |
| >=8 | 3 (37.5%) | 5 (83.3%) | 8 (57.1%) |
| Missing | 0 (0%) | 1 (16.7%) | 1 (7.1%) |
| **Baseline PSA** |  |  |  |
| Mean (SD) | 2.99 (2.56) | 7.11 (11.7) | 4.75 (7.81) |
| **Baseline PSA doubling time (months)** |  |  |  |
| Mean (SD) | 2.43 (1.06) | 3.51 (1.48) | 2.89 (1.33) |
| **Prior Treatment** |  |  |  |
| RT+ADT | 2 (25.0%) | 1 (16.7%) | 3 (21.4%) |
| Surgery+ADT | 2 (25.0%) | 1 (16.7%) | 3 (21.4%) |
| Surgery+RT | 3 (37.5%) | 0 (0%) | 3 (21.4%) |
| Surgery+RT+ADT | 1 (12.5%) | 4 (66.7%) | 5 (35.7%) |
| **RT to oligometastatic site(s)** |  |  |  |
| Yes | 2 (25.0%) | 4 (66.7%) | 6 (42.9%) |

### Efficacy

The disease progression-free rate at 12 weeks was 21.4% (MS-01, MS-05, MS-10). MS-01 had stabilization of PSA kinetics and received subsequent anti-cancer therapy 37.9 months after study initiation. MS-05 had initial stabilization of PSA kinetics, and at 9.6 months after study initiation, he was started on ATRA alone as the next line of therapy per local oncologist. He continues to experience stabilization of PSA without progression and has remained on ATRA alone for 6 years following study completion. MS-10 had a rising PSA on study without evidence of PCWG3-based PSA progression and was initiated on subsequent anti-cancer therapy 10.4 months after study initiation.

Among PSA-evaluated patients, the median PSADT was prolonged from 2.45 months at baseline to 4.56 months in the follow-up post-treatment phase **(Figure 1)**. Notably, all patients experienced testosterone recovery during the follow-up period, which was primarily attributed to the leuprolide washout and the prolongation of PSADT, resulting from the activity of AZA and ATRA. The median TTNT was 9.6 months, with four patients (28.6%) having a TTNT of ≥ 12 months. Two patients (14.3%) had a TTNT of more than 6 months. Of the total cohort, 5 (35.7%) patients experienced radiographic progression within 3 months of completing 5-AZA and ATRA. The treatment and follow-up course of each patient is described in **Figure 2**.

**Figure 1.**
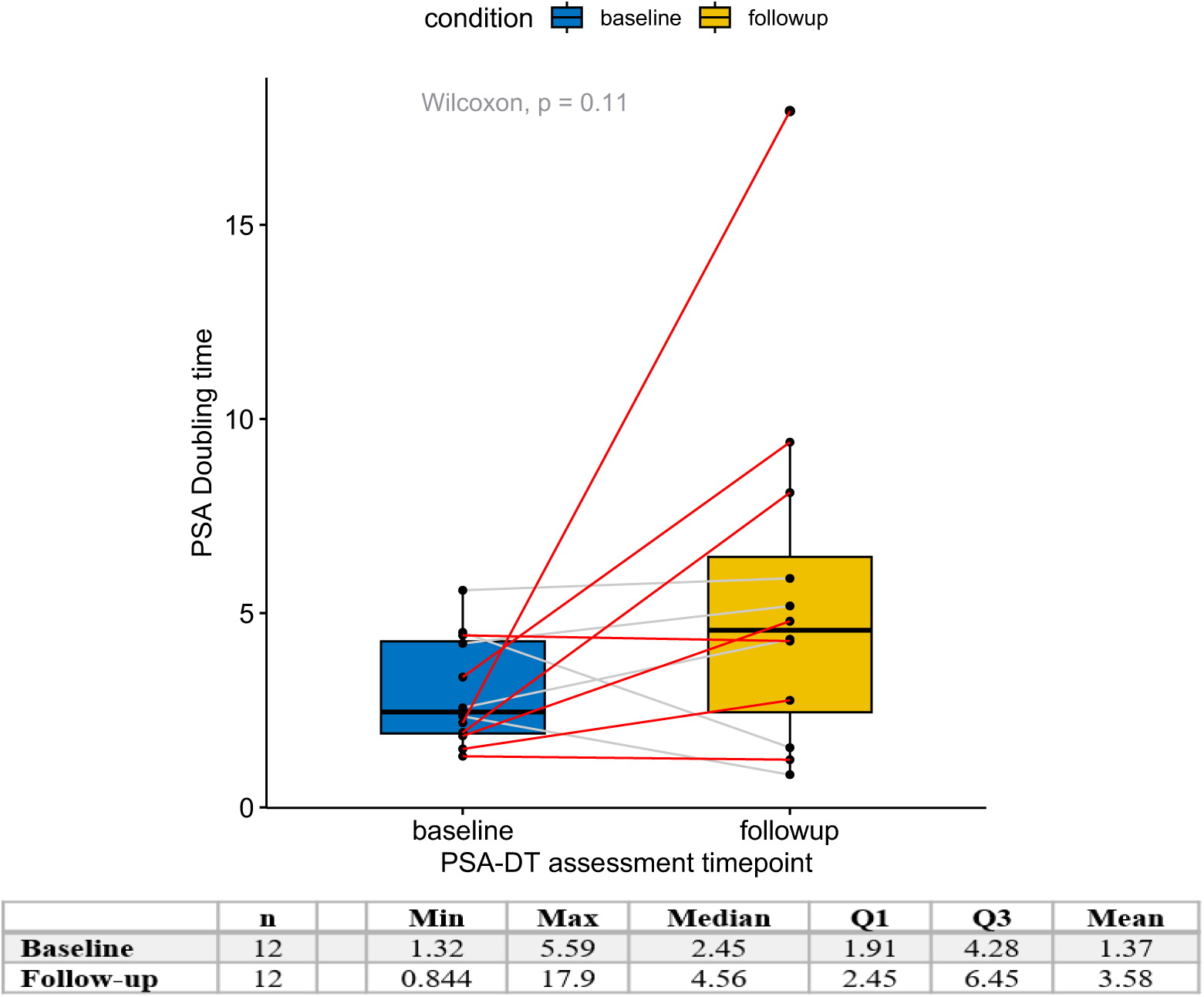
Change in PSA Doubling Time (PSADT) from Baseline to Follow-Up. The graph shows the change in PSA doubling time (PSADT) from baseline (blue) to follow-up (yellow). Each dot represents a patient, and the red line shows the change in PSADT from baseline to follow-up for the same patient. Time on the Y-axis is in months. Statistical test: Wilcoxon.

**Figure 2.**
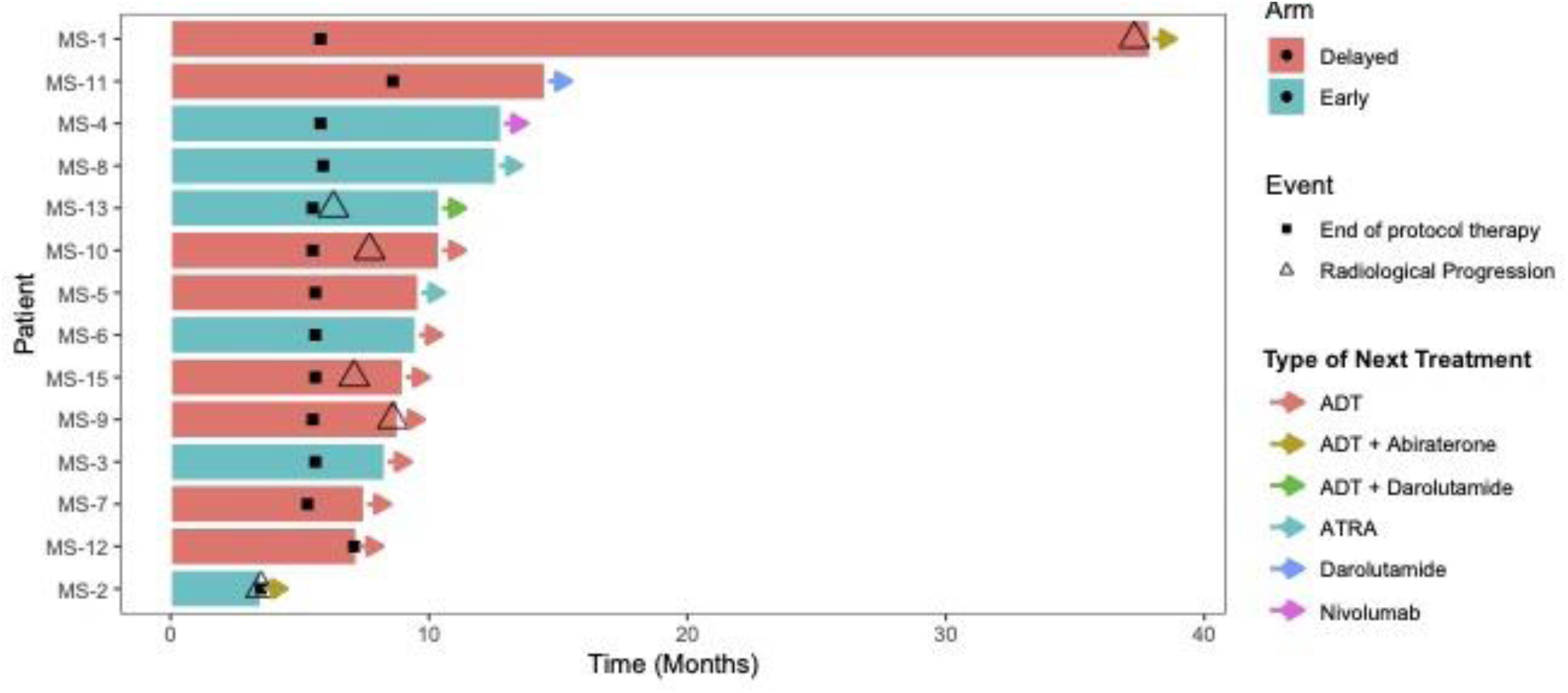
The swimmer’s plot shows the treatment and follow-up course of each patient.

### Safety

The incidence of treatment-emergent adverse events (TEAEs) and treatment-related adverse events (TRAEs) is shown in **Tables 2A and 2B**, respectively. The TEAEs were all grade 1 or 2, except for grade 3 hypertension and a decreased grade 3 lymphocyte count, which occurred in 1 patient each. Both were deemed to be non-treatment related. The most common TEAEs were anemia (57.1%), decreased lymphocyte count (35.7%), decreased white blood cell count (22.2%), and hyperglycemia (22.2%). The most common TRAEs were decreased white blood cell count (8.3%), hot flashes (8.3%), anemia (5.6%), and decreased lymphocyte count (5.6%). There were no dose interruptions, reductions, or discontinuations due to adverse events.

**Table 2a.** Incidence of treatment-emergent adverse events (TEAEs), by worst CTCAE grade for each patient.

| **AE term** | **Grade 1** | **Grade 2** | **Grade 3+** | **Total** |
| --- | --- | --- | --- | --- |
| **Anemia** | 6 (16.7) | 2 (5.6) | 0 | **8 (57.1)** |
| **Lymphocyte count decreased** | 1 (2.8) | 3 (8.3) | 1 (2.8) | **5 (35.7)** |
| **White blood cell decreased** | 6 (16.7) | 2 (5.6) | 0 | **8 (22.2)** |
| **Hyperglycemia** | 7 (19.4) | 1 (2.8) | 0 | **8 (22.2)** |
| **Aspartate aminotransferase increased** | 4 (11.1) | 0 | 0 | **4 (11.1)** |
| **Fatigue** | 4 (11.1) | 0 | 0 | **4 (11.1)** |
| **Hot flashes** | 4 (11.1) | 0 | 0 | **4 (11.1)** |
| **Hypertension** | 0 | 2 (5.6) | 1 (2.8) | **3 (8.3)** |
| **Urinary incontinence** | 1 (2.8) | 2 (5.6) | 0 | **3 (8.3)** |
| **Alanine aminotransferase increased** | 2 (5.6) | 0 | 0 | **2 (5.6)** |
| **Constipation** | 2 (5.6) | 0 | 0 | **2 (5.6)** |
| **Depression** | 2 (5.6) | 0 | 0 | **2 (5.6)** |
| **Gastroesophageal reflux disease** | 2 (5.6) | 0 | 0 | **2 (5.6)** |
| **Glaucoma** | 2 (5.6) | 0 | 0 | **2 (5.6)** |
| **Headaches** | 2 (5.6) | 0 | 0 | **2 (5.6)** |
| **Nausea** | 2 (5.6) | 0 | 0 | **2 (5.6)** |
| **Neutrophil count decreased** | 2 (5.6) | 0 | 0 | **2 (5.6)** |
| **Pain in extremity** | 2 (5.6) | 0 | 0 | **2 (5.6)** |
| **Platelet count decreased** | 2 (5.6) | 0 | 0 | **2 (5.6)** |
| **Pruritus** | 2 (5.6) | 0 | 0 | **2 (5.6)** |
| **Increased urinary frequency** | 2 (5.6) | 0 | 0 | **2 (5.6)** |
| **Atrial fibrillation** | 0 | 1 (2.8) | 0 | **1 (2.8)** |

**Table 2b.**
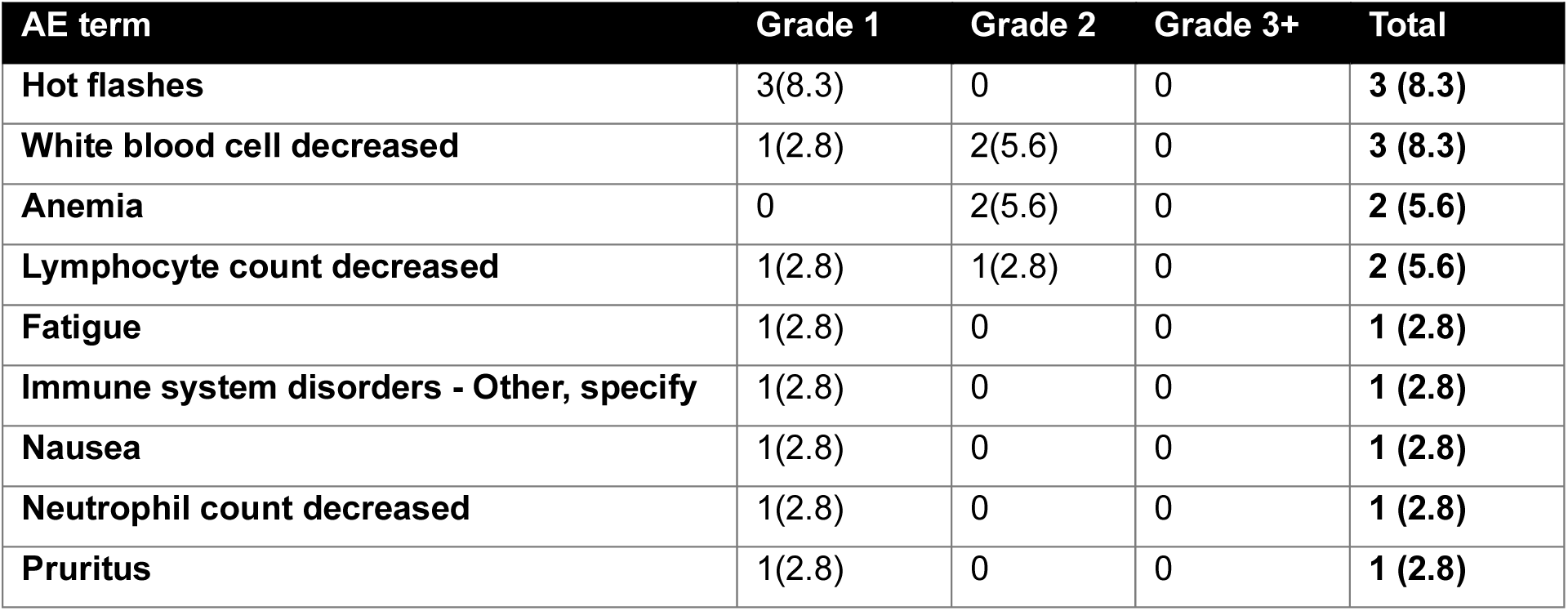
Incidence of treatment-related adverse events (TRAEs), by worst CTCAE grade for each patient.

### Dormancy Biomarkers

As a readout of the dormancy signature, we estimated the levels of BMP4 and BMP7 in the patient’s blood (4,7,23,24). Due to the unavailability of samples from patient MS-15, only thirteen patients were analyzed for dormancy biomarkers. Samples were collected at the following time points: before Lupron treatment *(Untreated)*, one month after the Lupron dose, and before the first dose of AZA + atRA *(Lupron)*, every three months after the first dose of AZA + atRA *(Treatment),* or monitoring cycle *(Monitoring),* and lastly during follow-ups *(Follow-up)* in both arms. To determine the baseline expression of BMP4 and BMP7 in the early and delayed arms, we quantified their expression at the untreated time point using ELISA. At baseline, the level of BMP4 in the early arm (early arm patients are in black text color) was undetectable in MS-02, MS-03, MS-08, and MS-13, and detected in MS-04 and MS-06 in the range of 66 to 127 pg/ml **(Fig. 3A)**. In the delayed arm (delayed arm patients are in green text color), the level of BMP4 was undetectable in MS-01 and MS-05. In patients MS-07, MS-09, MS-10, MS-11, and MS-12, the levels detected ranged from 18 to 110 pg/mL **(Fig. 3A)**. The BMP4 level in the early arm did not increase significantly with AZA + atRA treatment. Patients MS-04 and MS-06 maintained BMP4 levels until the first follow-up and last monitoring cycle, respectively, after which they dropped to an undetectable level **(Fig. 3A)**. In the delayed arm, patients MS-01, MS-05, and MS-10 were responders (curves are in blue color) to AZA + atRA treatments based on clinical outcomes **(Fig. 3A)**. MS-05 showed a significant increase in BMP4 level (up to ∼250 pg/ml) upon treatment and maintained this level until the penultimate AZA + atRA cycle. The baseline BMP4 level in MS-10 was already high, and with treatment, it maintained this level until the penultimate follow-up cycle, after which it dropped to an undetectable level. Interestingly, the BMP4 level in MS-01 did not increase until the last treatment cycle, but it eventually rose during follow-ups **(Fig. 3A)**. The baseline BMP7 levels were undetectable in MS-02, MS-03, MS-01, and MS-05, while in patients MS-04, MS-06, MS-08, MS-13, MS-07, MS-09, MS-10, MS-11, and MS-12, the BMP7 levels ranged from 82 to 922 pg/ml **(Fig. 3B)**. Patients MS-06, MS-08, and MS-13 in the early arm consistently maintained similar levels of BMP7 compared to the baseline before dropping to an undetectable level in follow-ups **(Fig. 3B)**. In the delayed arm, MS-05 and MS-10, which were responders, showed an overall increase in BMP7 levels, and their levels were maintained until the second follow-up. Similar to BMP4, BMP7 also increased during follow-ups in MS-01 **(Fig. 3B)**.

**Figure 3.**
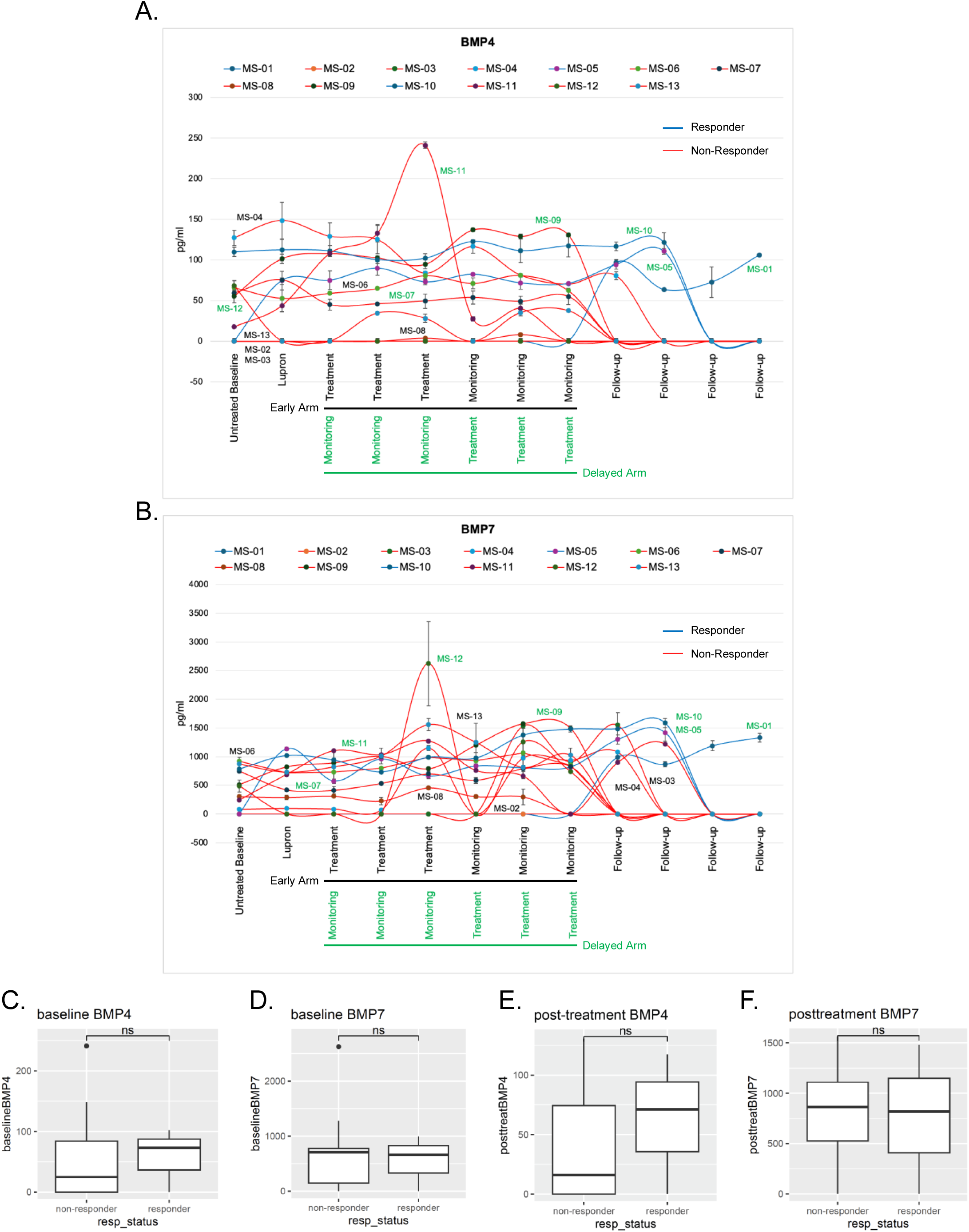
Change in the BMP4 and BMP7 cytokine levels in the blood of patients during treatment. **A-B.** The amount of BMP4 and BMP7 in the blood was measured by ELISA in technical duplicates. Clinically responder and non-responder patients are shown in blue and red lines. Patients in the early arm and the delayed arm are labeled in black and green fonts. **C-F.** Graphs showing the cumulative levels (pg/ml) of BMP4 and BMP7 in responders vs non-responders at the baseline and post-treatment stage. In the post-treatment, all the measurements were considered from Lupron to follow-ups. Statistical test: Wilcoxon.

Furthermore, we compared the levels of BMP4 and BMP7 from all the non-responder patients (MS-02, MS-03, MS-04, MS-06, MS-07, MS-08, MS-09, MS-11, MS-12, and MS-13) to those of the responder patients (MS-01, MS-05, and MS-10) **(Fig. 3C-F)**. At baseline (untreated), BMP4 levels were relatively high in responders, and no difference was observed in BMP7 levels between responders and non-responders **(Fig. 3C-D)**. Similarly, responders displayed relatively higher BMP4 levels but no difference in BMP7 levels post-treatment **(Fig. 3E-F)**. Although the differences in BMP4 levels between responders and non-responders were statistically not significant, they suggest that maintaining relatively high levels of both BMP4 and BMP7 over time serves as a good prognostic marker.

Next, we examined the effect of AZA + atRA treatment on metastatic load by analyzing the number of CTCs in the blood. We detected CTCs (processed from 10 ml of blood) using immunofluorescence staining of pan-cytokeratin (Pan-CK+ CTC), CD45-(contaminating blood cells), and NR2F1+ (dormancy marker expressed by putative dormant prostate cancer cells **(Fig. 4A)** (9). At the untreated baseline time point, patients in the early arm showed a relatively higher frequency of CTCs (0 to 9) compared to the delayed arm, where CTCs were not detected in most patients **(Table 3)**. Only MS-11 and MS-12 had 4 and 1 CTCs, respectively. Later during treatment, most patients in the early arm showed a very low frequency of CTCs (0 to 4). On the other hand, patients in the delayed arm showed a relatively higher number of CTCs; however, many of these were NR2F1 positive, as seen in MS-01, MS-11, and MS-12 **(Table 3)**. In responder patients MS-05 and MS-10, we only detected CTCs at two and one time points, respectively. In MS-01, we detected a relatively high number of CTCs, with 50% or more being NR2F1 positive. A comparison of CTCs at baseline between responder and non-responder patients revealed a relatively low number of CTCs in responder patients **(Fig. 4B)**. Post-treatment, the frequency of CTCs in responders was low compared to non-responders, but the difference was statistically not significant **(Fig. 4C)**.

**Figure 4.**
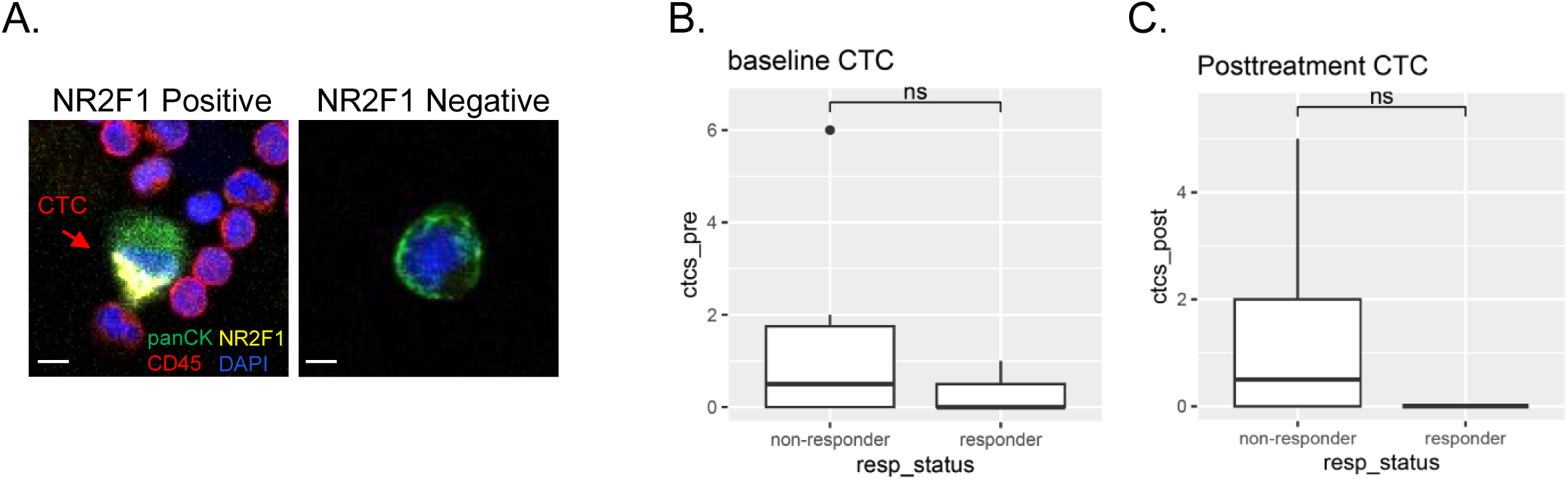
CTC counts in responder vs non-responder patients. **A.** Immunofluorescence staining of CTCs detected in patients’ blood samples. CTCs are stained with pan-cytokeratin (green), NR2F1 (yellow), and blood cells are positive for CD45 (red). DAPI was used to detect the nucleus. **B-C.** Average count of CTCs in responder and non-responder patients at the baseline and post-treatment stage. Due to the low number of NR2F1-positive CTCs, we excluded them from the analysis. Statistical test: Wilcoxon.

**Table 3.** CTC counts.

| Early Arm |  |  |  |  |  |  | Delayed Arm |  |  |  |  |  |  |  |
| --- | --- | --- | --- | --- | --- | --- | --- | --- | --- | --- | --- | --- | --- | --- |
|  | MS-02 | MS-03 | MS-04 | MS-06 | MS-08 | MS-13 |  | MS-01 | MS-05 | MS-07 | MS-09 | MS-10 | MS-11 | MS-12 |
| Untreated Baseline | 7 | 6(1) | 9(4) | UD | 1 | UD | Untreated Baseline | UD | UD | UD | UD | UD | 4 | 1(1) |
| Lupron | UD | 1 | 1 | UD | UD | 2 | Lupron | SNA | UD | UD | UD | UD | 1 | SNA |
| Treatment | SNA | SNA | 1 | UD | UD | 1 | Monitoring | SNA | UD | UD | UD | 5 | UD | SNA |
| Treatment | SNA | SNA | UD | UD | UD | 3 | Monitoring | SNA | UD | UD | UD | UD | UD | SNA |
| Treatment | 5 | 5(3) | UD | UD | 1 | 2 | Monitoring | 3(2) | UD | UD | 2 | UD | 10 (4) | UD |
| Monitoring | SNA | UD | 1 | 1 | UD | 4 | Treatment | 11 | UD | UD | 2 | UD | 2(2) | UD |
| Monitoring | SNA | UD | UD | 1 | UD | 1 | Treatment | 6(3) | 4 | UD | 1 | UD | 1(1) | 1(1) |
| Monitoring | SNA | 1 | UD | UD | SNA | UD | Treatment | 3(3) | UD | UD | 4 | UD | SNA | 1(1) |
| Follow-up | SNA | UD | 1 | SNA | SNA | SNA | Follow-up | UD | 1 | SNA | SNA | UD | 1(1) | SNA |
| Follow-up | SNA | SNA | SNA | SNA | SNA | SNA | Follow-up | UD | UD | SNA | SNA | UD | 3(1) | SNA |
| Follow-up | SNA | SNA | SNA | SNA | SNA | SNA | Follow-up | 1 (1) | SNA | SNA | SNA | SNA | SNA | SNA |
| Follow-up | SNA | SNA | SNA | SNA | SNA | SNA | Follow-up | 1(1) | SNA | SNA | SNA | SNA | SNA | SNA |

( ): NR2F1 positive CTCs; UD: Undetected; SNA: Sample Not Available; patients in green font are responders.

## Discussion

To our knowledge, this is the first clinical trial to evaluate epigenetic reprogramming as a therapeutic strategy to modulate tumor dormancy pathways in biochemically recurrent prostate cancer (4,5,9,13,17). Our findings demonstrate that the combination of 5-Azacitidine (AZA) and all-trans retinoic acid (ATRA) was well tolerated, with no dose-limiting toxicities or treatment discontinuations due to adverse events. Although the observed disease progression-free rate at 12 weeks was modest (21.4%), the median PSA doubling time (PSADT) was prolonged after treatment by 2 months, suggesting a long-lived biological effect following a 3-month treatment period. Notably, four patients exhibited a prolonged time to next treatment (≥12 months), indicating a potential clinical benefit in select patients of a relatively short treatment cycle. However, the early radiographic progression observed in 5 of 14 patients underscores the need for further refinement of this approach, including patient selection and identification of predictive biomarkers of dormancy response. NR2F1 detection in primary lesions may inform on potential responders depending on the NR2F1 status.

The biology of tumor dormancy is increasingly recognized as a critical mechanism of disease progression in solid tumors, including prostate cancer. DTCs can persist in a non-proliferative state for years, evading immune surveillance and conventional therapies such as ADT. Preclinical studies have implicated epigenetic mechanisms—particularly DNA methylation and chromatin remodeling—in maintaining dormancy and suppressing reactivation (9,11). Agents such as AZA have been shown to restore tumor-suppressive gene expression programs associated with dormancy (9,16). while ATRA has been associated with cellular differentiation and suppression of metastatic progression (9). NR2F1 agonists have also been tested as drugs to induce cancer cell dormancy and prevent metastasis (25). Based on these findings, we hypothesized that the combination of AZA and ATRA could promote dormancy in DTCs and delay disease progression in biochemically recurrent PCa.

For patients with biochemically recurrent prostate cancer, ADT remains the mainstay, either as intermittent or continuous therapy. While ADT effectively suppresses PSA levels and delays metastases, it is associated with significant toxicities, including reduced quality of life, cardiovascular risk, bone loss and fractures, and metabolic dysfunction. Recent trials have evaluated intensified hormonal approaches, such as adding novel androgen receptor pathway inhibitors (e.g., enzalutamide) to ADT, demonstrating improvements in metastasis-free survival (26). However, these regimens come with added toxicity and are only approved for those with biochemical recurrence at “high risk” of developing metastases.

Our study presents a distinct paradigm; rather than suppressing PCa growth through androgen blockade, we aimed to modulate the epigenetic landscape of DTCs to promote dormancy. If validated in larger trials, this approach could delay disease progression while minimizing the morbidity of continuous hormonal suppression. Patients with slow PSADT may particularly benefit from dormancy-inducing therapies as an alternative to immediate ADT initiation.

One patient (MS-05), who received ATRA monotherapy after completion of the study treatment(s) as a “maintenance therapy,” has had stabilization of PSA for over 6 years. The reason for prolonged clinical activity is unclear; however, it is postulated that ATRA may extend dormancy signaling as monotherapy after initial induction of dormancy by AZA-ATRA combination. ATRA has been shown to induce remission in various cancers, and responders to this disease were documented across cancers and experimental models (27–29). However, further work is needed to elucidate the role of ATRA monotherapy in maintaining dormancy durably.

Although the effect of AZA-ATRA combination therapy on dormant biomarkers was not statistically significant due to the small number of patients, the increased pro-dormancy cytokine (BMP4 and BMP7) levels in MS-01, MS-5, and MS-10 suggest an association with better prognosis and increased survival. Our data suggest that patients who already have relatively higher BMP4 and BMP7 expression levels and maintain them due to AZA-ATRA treatment throughout the study period showed better survival. Interestingly, we observed a higher number of CTCs in MS-01, MS-05 and MS-10 patients; however, with treatment, we saw a decrease in the CTC count, as well as an increase in NR2F1-positive CTCs. Previously, we have shown that AZA-ATRA treatment increases NR2F1 expression (9).

Despite our initial evidence, several limitations of our study warrant consideration. The small sample size and single-institution design limit generalizability, and the observed heterogeneity in response suggests that patient selection may be critical for optimizing therapeutic benefit. The biomarker analysis in the blood samples of patients provided strong cues about the pro-dormancy initiation by the AZA-ATRA treatment and their role in patient’s survival. However, due to the low number of patients and the limited granularity of analysis, additional analysis may provide insight into the mechanisms of response and resistance. Additionally, the durability of the dormancy effect remains a key question. While our data suggest an impact on prolonging PSADT in a subset of patients, longer follow-up is needed to assess whether dormancy induction translates to meaningful delays in metastatic progression. It is also worth considering whether repeating the treatment may enhance efficacy. Lastly, the use of leuprolide at the beginning of the treatment course may have confounded the findings when assessing response in the two treatment arms. Future studies should explore alternative dosing strategies, including a defined washout period between ADT and dormancy reprogramming therapy.

### Conclusion

This study represents the initial clinical exploration of epigenetic reprogramming as a therapeutic approach for biochemically recurrent prostate cancer. While preliminary signals of efficacy were observed, further research is needed to refine patient selection criteria, identify dormancy biomarkers, and optimize the treatment schedule.

## Authors’ Contributions

V.P. – resources, data curation, investigation, methodology, analysis, writing, and editing. D.K.S. – methodology, investigation, data curation, visualization, analysis, writing, and editing. H.J. – analysis, and writing. N.S. – methodology. B.L. – investigation. C-K.T. – investigation. M.D.G. – investigation. L.D. – investigation. M.L-A. – methodology. M.S.S. – conceptualization and editing. J.A.A-G. – conceptualization, funding acquisition, resources, supervision, writing, and editing. W.K.O. - conceptualization, resources, supervision, writing, and editing.

## Data Availability

All data produced in the present study are available upon reasonable request to the authors

## Acknowledgments

This work has been supported by the National Institute of Health (NIH, National Clinical Trial ID: NCT03572387), Jimmy V Foundation (T2016-008), NIH/National Cancer Institute (NCI) CA109182-23 to J.A.A-G.

---

**Table S1.**
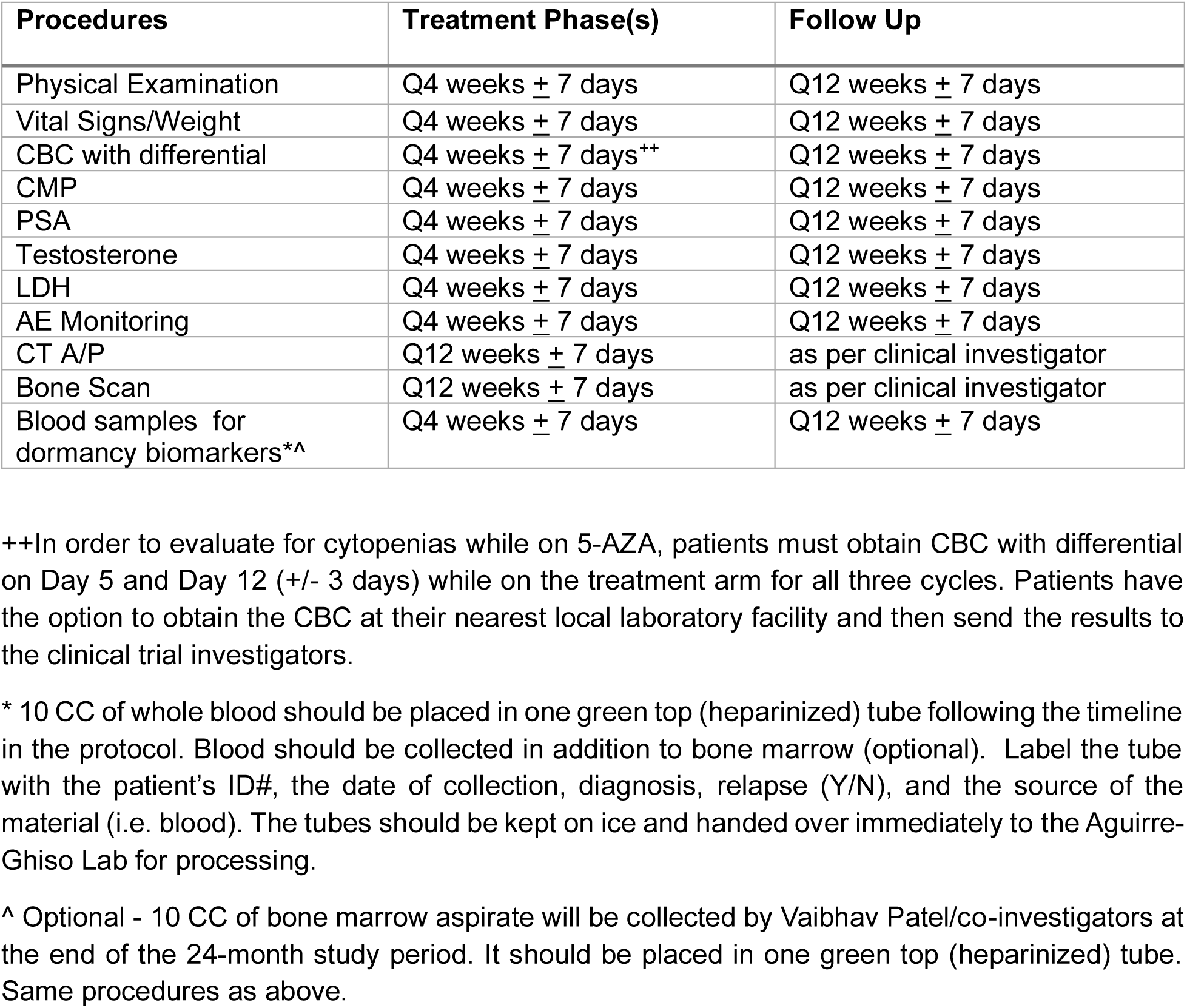
On-study Schedule of Activities.

